# Assessment of small pulmonary blood vessels in COVID-19 patients using HRCT

**DOI:** 10.1101/2020.05.22.20108084

**Authors:** Muriel Lins, Jan Vandevenne, Muhunthan Thillai, Ben R. Lavon, Maarten Lanclus, Stijn Bonte, Rik Godon, Irvin Kendall, Jan De Backer, Wilfried De Backer

## Abstract

**Background:** Mounting evidence supports the role of pulmonary hemodynamic alternations in the pathogenesis of COVID-19. Previous studies have demonstrated that changes in pulmonary blood volumes measured on CT are associated with histopathological markers of pulmonary vascular pruning, suggesting that quantitative HRCT analysis may eventually be useful in the assessment pulmonary vascular dysfunction more broadly.

**Methods:** Building upon previous work, automated HRCT measures of small blood vessel volume and pulmonary vascular density were developed. Scans from 103 COVID-19 patients and 108 healthy volunteers were analyzed and their results compared, with comparisons made both on lobar and global levels.

**Results:** Compared to healthy volunteers, COVID-19 patients showed significant reduction in BV5 (pulmonary blood volume contained in blood vessels of <5 mm^2^) expressed as BV5/(Total pulmonary blood volume) (p<0.0001), and significant increases in BV5_10 and BV 10 (pulmonary blood volumes contained in vessels between 5 and 10 mm^2^ and above 10 mm^2^, respectively) (p<0.0001). These changes were consistent across lobes.

**Conclusions:** COVID-19 patients display striking anomalies in the distribution of blood volume within the pulmonary vascular tree, consistent with increased pulmonary vasculature resistance in the pulmonary vessels below the resolution of HRCT.

## Introduction

In December 2019 a novel beta coronavirus, since dubbed SARS-CoV2, emerged as the cause of an outbreak of respiratory disease beginning in Wuhan, China, before eventually spreading widely across the globe. The resulting disease, named COVID-19 by the World Health Organization, remains mild or asymptomatic in the majority of those infected, but causes severe or potentially life-threatening disease in a minority, most of whom are thought to have preexisting comorbidities. Initially it was believed that severe refractory hypoxia and death in COVID-19 patients was the result of viral pneumonia progressing to acute lung injury and, in severe cases, Acute Respiratory Distress Syndrome (ARDS). As the pandemic progressed, a distinct clinical picture began to emerge of patients with progressive hypoxia but relatively well-preserved lung function and compliance which is inconsistent with conventional understandings of ARDS [1]. Alongside emerging understanding of the role of the ACE2 pathway (the entry receptor for the virus) in the pathogenic process, it has become clear that at least some COVID-19 patients exhibit significant pulmonary vascular involvement [2, 3]. While the nature of this involvement remains unclear, widespread coagulopathy, microthrombi, and dysregulated hypoxic pulmonary vasoconstriction (HPV) have all been implicated [1, 3].

In previous work examining pulmonary vascular remodeling in smokers, significant correlations have been drawn between histopathological markers of microvascular pruning and CT-derived markers of pulmonary blood loss in the smallest caliber vessels that may be resolved on scans [4]. These results suggest that while the pathological changes to the pulmonary vasculature (“pulmonary vascular remodeling”, e.g. intima thickening, increased muscularization and basal tone of pulmonary arterial smooth muscle) implicated in pulmonary vascular disease (PVD) occur primarily below the resolution of HRCT scans, those processes do have a measurable effect on larger, more proximal vessels. While the clinical significance of these quantities is not yet clear, they hold promise as potential noninvasive measures of pulmonary hemodynamics that may be valuable in the development of more effective diagnosis and treatment of PVD.

In this study we assessed the use of novel HRCT-derived measures of pulmonary blood volume and pulmonary vascular density in patients with COVID-19. We hypothesized that the so-called “silent hypoxia” noted in hospitalized patients may be due impaired gas exchange resulting from abnormalities in the distribution of pulmonary blood volumes, and moreover that HRCT may offer clues as to the processes occurring in pulmonary arterioles and capillaries.

## Materials and Methods

103 hospitalized COVID-19 patients were retrospectively selected based on the availability of HRCT scans of the lungs with slice thickness 1.5 mm or less, as well as PCR-confirmed SARS-CoV2 infection. HRCT scans were provided by the respective hospitals where they were acquired. Institutional Review Board approval was granted by the respective local committees, and written informed consent was obtained. 83 scans came from Belgium (43 from AZ Sint-Maarten (Mechelen), 40 from Ziekenhuis Oost-Limburg (Genk)), 10 from the UK (Royal Papworth Hospital) and 10 from China (Wenzhou Medical University). 108 inspiratory scans from healthy patients were acquired from the COPDGene cohort (NCT00608764). Because the patient scans were acquired in the course of clinical care and without a standardized protocol, slice thickness varied between .6 mm and 1.5 mm. The demographics for these cohorts are shown below in (Table 1).

**Table 1.** Demographics.

|  | COVID-19 patients (N=103) | Healthy patients (N=108) |
| --- | --- | --- |
| Mean age (SD), years | 58.08 (12.29) | 62.3 (9.14) |
| Female/Male/Unknown | 17/36/50 | 73/35/0 |
| Mean height (SD), cm | 173.6 (9.34) | 166.4 (9.3) |

SD, standard deviation

3D reconstructions of the lungs and pulmonary vasculature were created. An automated blood vessel segmentation algorithm performs an eigenvalue analysis of the Hessian matrix to enhance and identify tubular structures, by returning the probability of each voxel belonging to tubular structure based on shape analysis [5]. Next, Hounsfield unit (HU) thresholds are used to limit the vessels. The HU thresholds are based on the vessels size and are defined by an automated adaptive iterative threshold method. In the preprocessing, a gradient anisotropic diffusion filter is applied, and a region of interest is defined to remove some false positives. Subsequently, the smaller non-connected parts are removed. To account for the effects of slice thickness on results, sensitivity analysis was performed. Volumes were then computed from the cross-sectional area of each vessel. Following the convention established by Rahagi et al, the volume of blood contained in vessels below 5 mm^2^ cross sectional area (down to a cutoff of 1.25 mm^2^) was termed “BV5”. We additionally defined BV5-10 as the volume of blood contained in vessels with cross-sectional area between 5 and 10 mm^2^, and BV10 as the volume contained in vessels with cross-sectional area above 10 mm^2^. We refer collectively to these quantities as BVX.

To account for variation in lung volume, we chose to normalize BVX by total pulmonary blood volume. This permits for the computation of a “BV spectrum”, a curve representing the percent of total pulmonary blood volume contained within vessels of a given caliber as a function of cross-sectional area (Figure 1). It had previously been observed in analysis of scans from healthy volunteers that this yielded values with very low variance over all scales (Figure 1)

**Figure 1:**
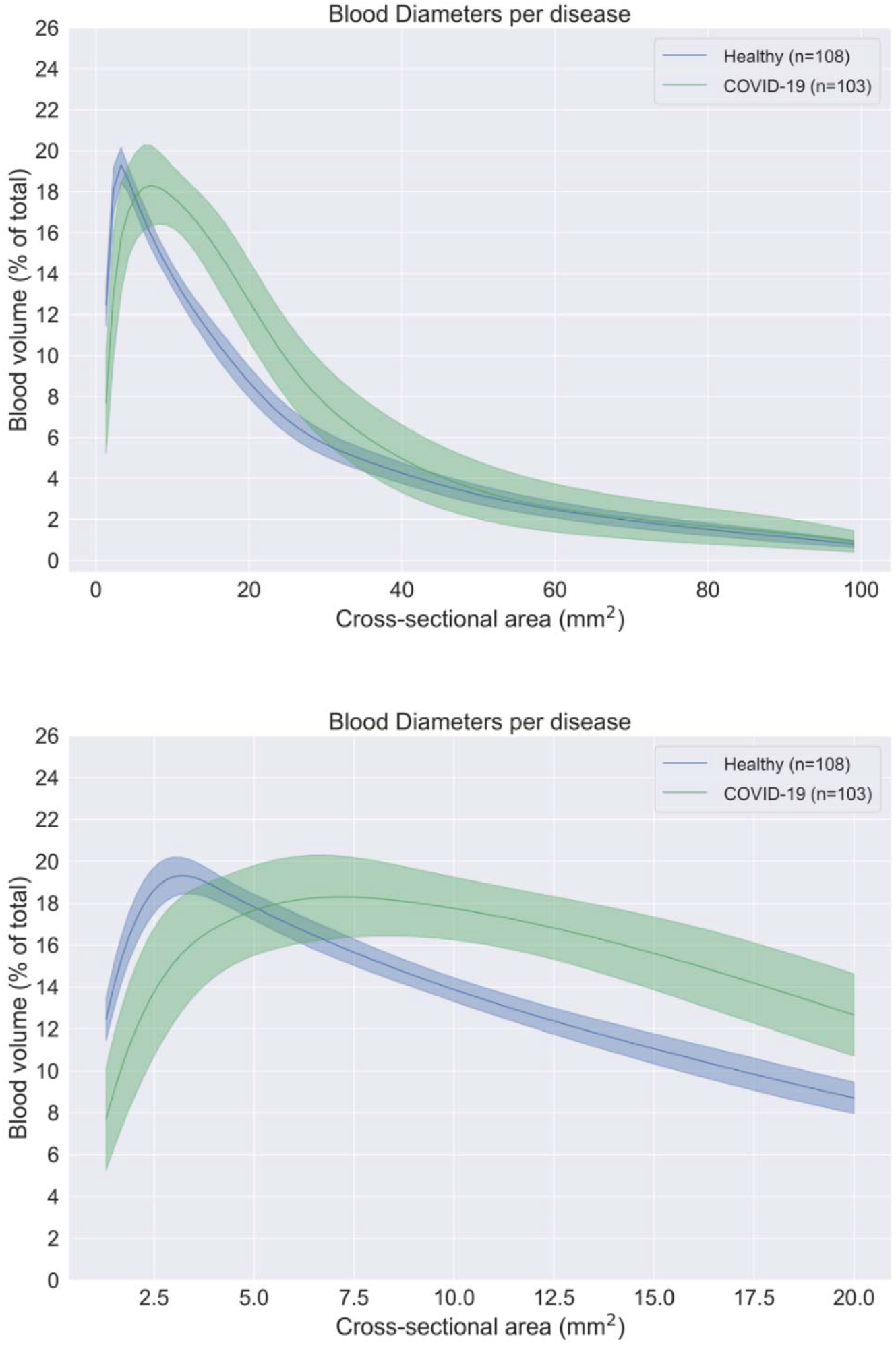
Spectrum plot describing the blood volume as a function of the blood vessel cross-sectional area for the large spectrum (top) reflecting most blood vessels visible in the HRCT scans and the small spectrum (bottom) zooming in on the small blood vessels visible in the HRCT scans.

COVID-19 patients often present with various opacities on CT. Because these regions are more dense than surrounding tissue, they may be confused for blood by our segmentation algorithm (and vice versa). In an effort to control for this and to understand the scope of its impact on our results, a fully convolutional deep learning model was trained on a set of manually labeled axial CT slices to detect and segment ground glass opacities, reticulation, and areas of consolidation, and estimate the volume of affected tissue.

Two-sample t-tests were used to assess significance (p < 0.05). All analyses were performed using the open-source statistical environment R version 3.2.5 or higher (The R Foundation for Statistical Computing, Vienna, Austria).

## Results

The results of BVX analysis can be seen in Figure 2, below. Compared with healthy volunteers, patients from the COVID-19 cohort have markedly reduced BV5 (p<.0001), with corresponding increases in BV5-10 (p<0.0001) and BV10 (p<0.0001),

**Figure 2:**
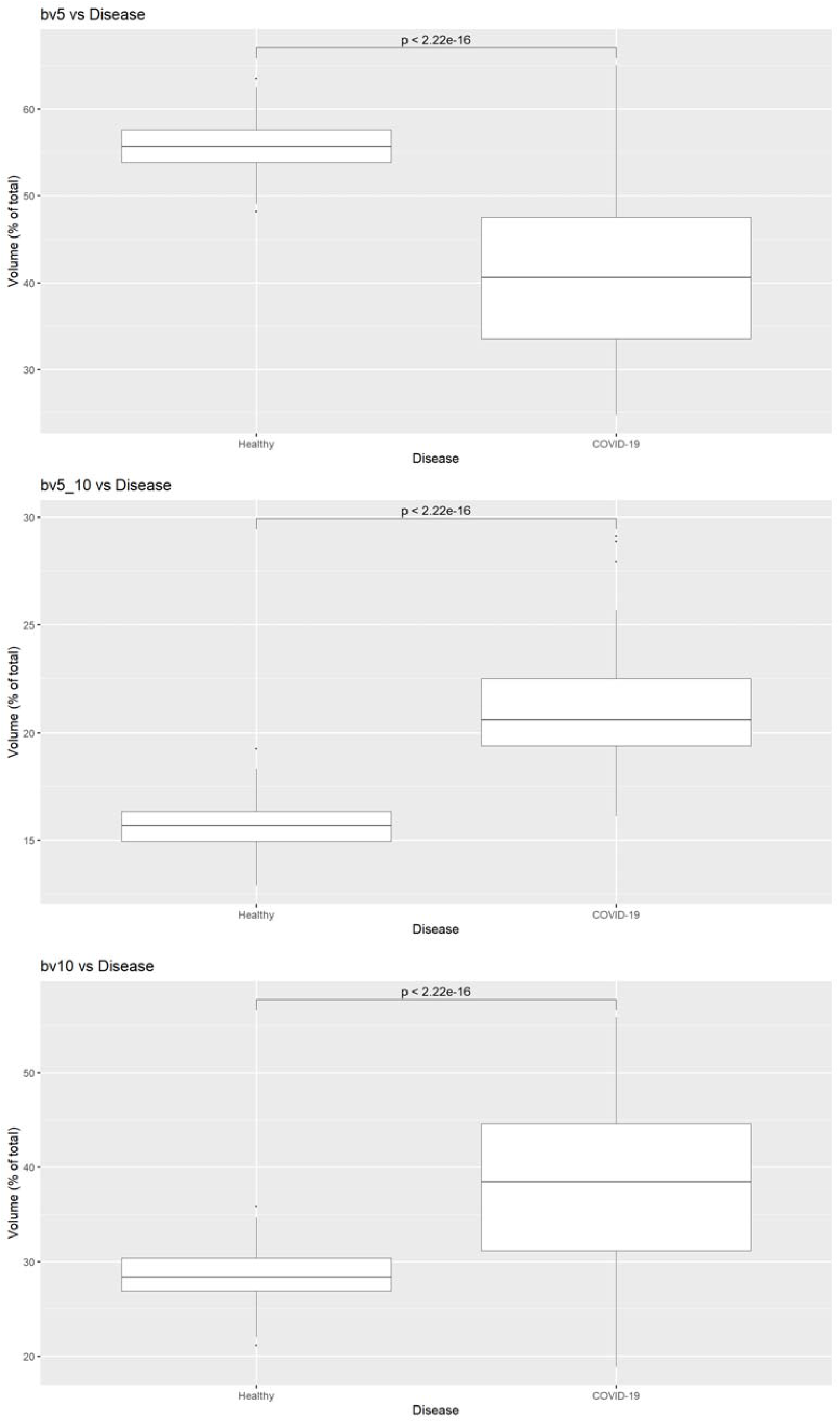
Comparison of BV5, BV5-10, and BV10 (expressed as % total pulmonary blood volume) between healthy volunteers and patients with COVID-19

**Figure 3:**
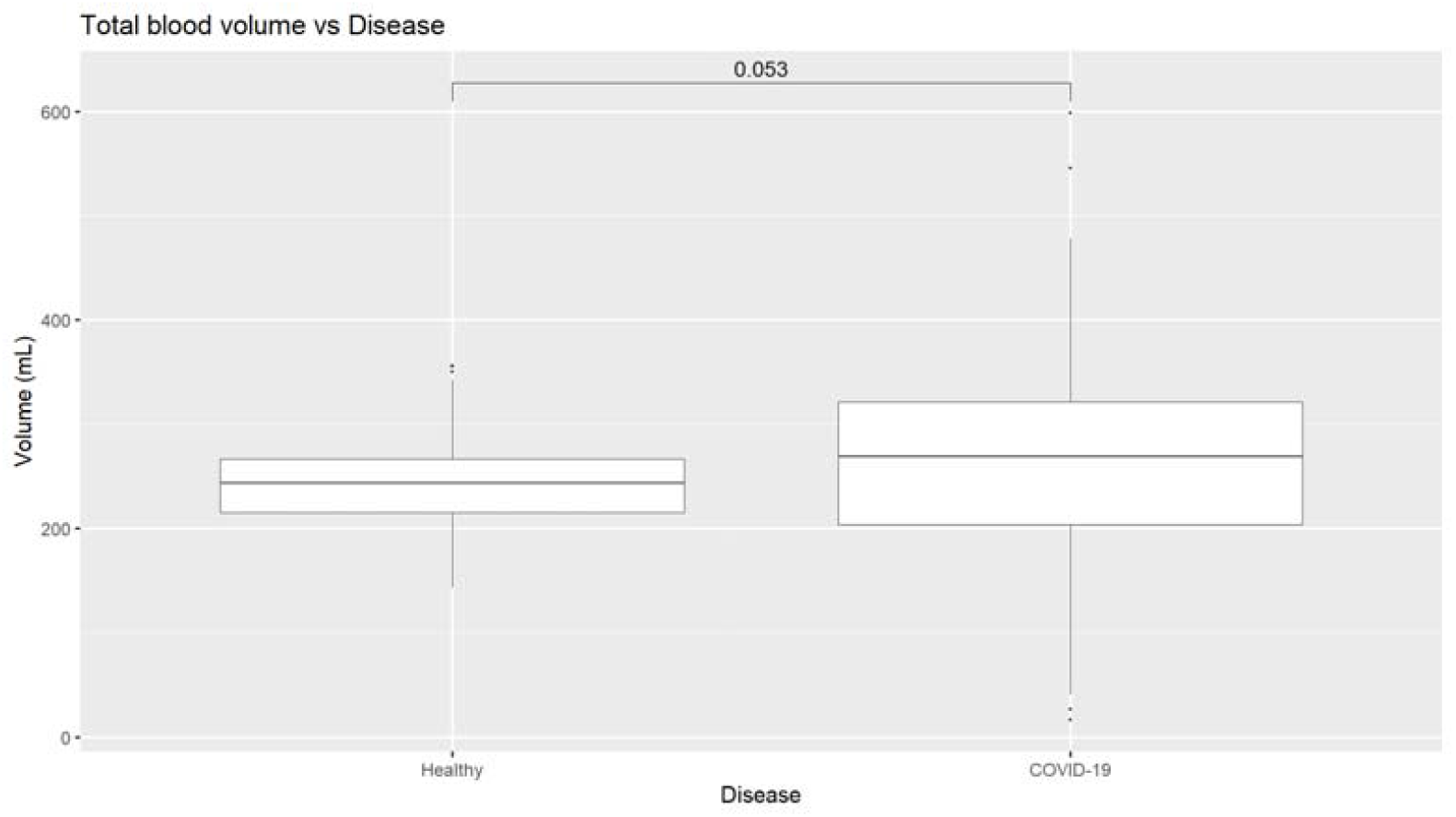
Comparison of total pulmonary blood volume between healthy volunteers and patients with COVID-19.

Figure 1 shows the BV spectra for the COVID-19 and healthy cohorts overlaid, and allows for a qualitative grasp of this redistribution. The COVID-19 curve dips below the healthy curve at approximately 5mm^2^, with a flatter-than-usual peak to the right of the healthy patients at just below 7.5 mm^2^, and remaining elevated until ∼50mm^2^ before normalizing. This represents a sharp drop in the amount of blood contained in small vessels with a more gradual renormalization over the medium and larger sized vessels. These changes can be seen quite strikingly in Figure 4, which compares 3D reconstructions of the pulmonary vasculature of a representative healthy and COVID-19 patient. Segments are color-coded according to size, and the relative and marked absence of small vessels (colored red) is notable, as is the marked “proliferation” of medium-sized yellow vessels.

**Figure 4:**
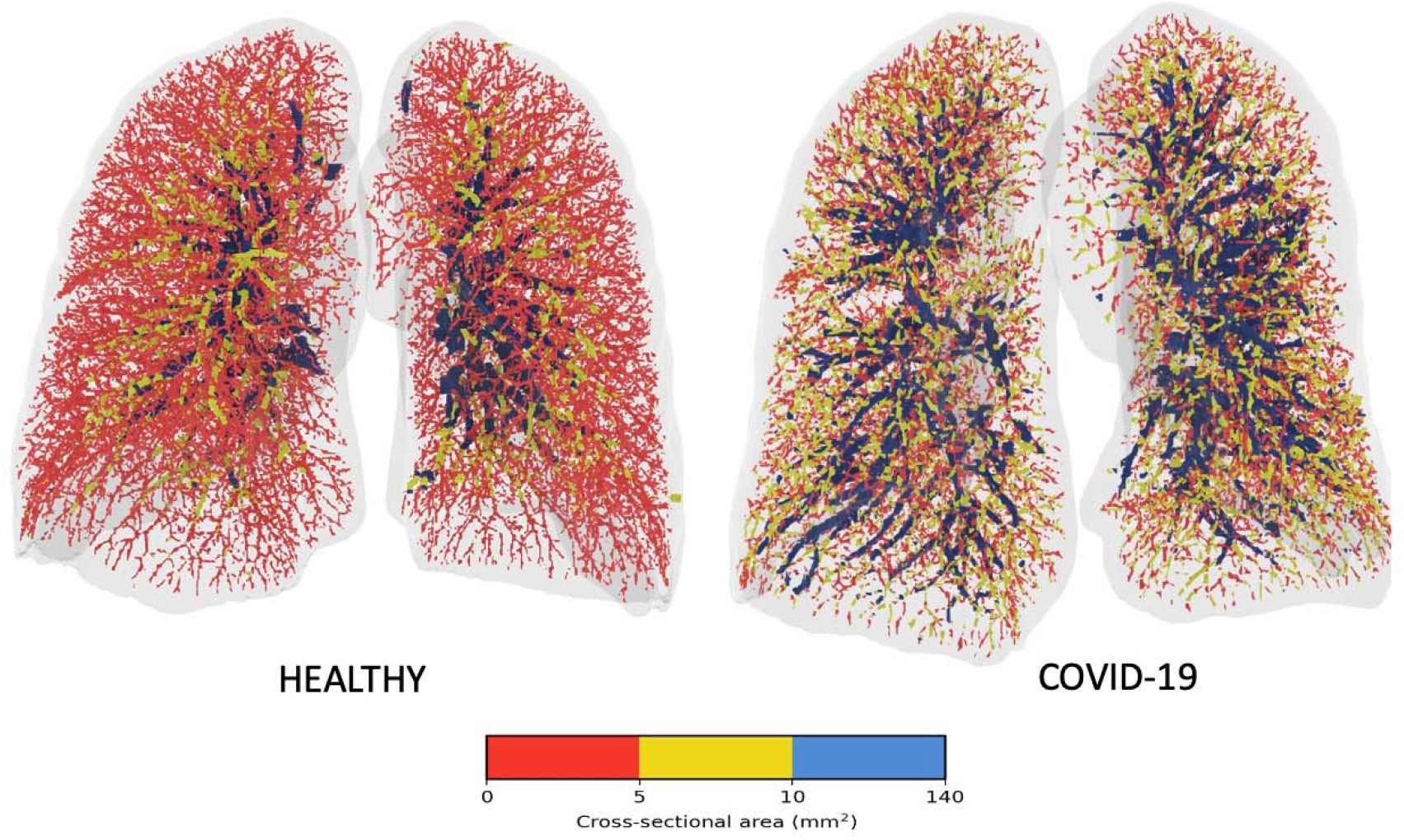
Visual representation of the blood vessels colored according to their size. Red denotes the small vessels, yellow the mid-size vessels and blue the larger vessels.

Figure 5 shows the results of a mixed effect model of the relationship between BVX and the opacity score on a lobar level. BV5 shows a significant but modestly inverse relationship with increasing opacities (p<0.01, R^2^ = 0.18). BV5-10 was likewise inversely related, but the effect size was even smaller (p= 0.01, R^2^ = 0.02), and BV10 shows a direct relation with opacity (p<0.01, R^2^ =0.21).

**Figure 5:**
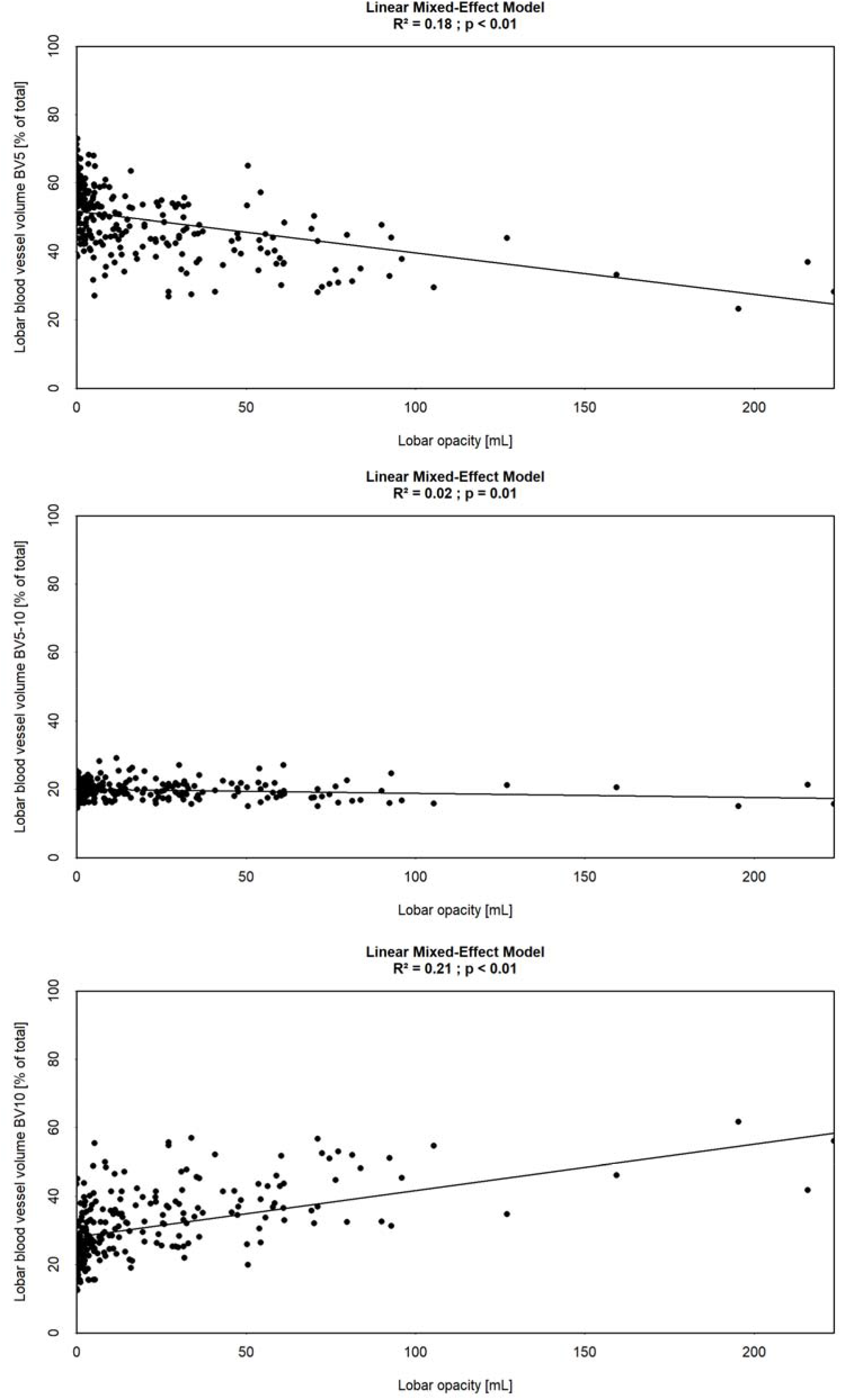
Mixed effect model BV5 (top), BV5-10 (middle), and BV10 (bottom) as function of lobar opacity (ml), assessed by lobe

## Discussion

While it remains unclear how SARS-CoV2 infection leads to the constellation of clinical and imaging signs associated with COVID-19, certain elements are well established. The virus has been discovered to enter cells by way of the angiotensin-converting enzyme 2 (ACE2) receptor, and further to downregulate its expression [2]. Because ACE2 interfaces with the vasoactive renin-angiotensin-aldosterone system (RAAS), this pathway has been implicated as the cause of vascular anomalies observed in COVID-19 patients [2].

Because total pulmonary blood volumes as measured on CT does not vary significantly between groups (Figure 3), this loss of blood volume in small vessels and increase of blood volumes in medium and large vessels can be interpreted as a “redistribution” of blood throughout the pulmonary vasculature, in particular from smaller vessels to larger vessels.

The precise mechanism leading to this redistribution remains unknown, but these results may point towards a better understanding of the pathophysiology involved. The observed effects could be explained by increased pulmonary vascular resistance (PVR) downstream which, given constant cardiac output and left atrial pressure, is concomitant with increased pulmonary arterial pressure. This could result from dysregulated vasoconstriction leading to increased tone in the pulmonary arteries, or by occlusion due to microthrombi or tissue damage. This would be consistent with the observed pattern of hypoxia in the context of preserved lung compliance, with SAR2-CoV2 infection leading to an infectious mimic of idiopathic pulmonary hypertension, which is associated with both coagulopathy and increased muscularization of pulmonary arteries [6]. Future studies could elucidate these mechanisms further by way of more careful analysis of changes in BVX, additional hemodynamic data from COVID-19 patients, or potentially through pathological examination of lung tissue.

It should be noted that in most anatomical studies of human pulmonary vasculature the “muscularized” pulmonary arteries, which are thought to be primarily (but not exclusively) responsible the hypoxic pulmonary vasoconstriction, have diameters in the range of 70-1000 µm [7-10]. This range corresponds to cross-sectional areas of ∼.004-.79 mm^2^, somewhat below the resolution of current HRCT. We quantify vessels as small as 1.25 mm^2^ ; vessels this size and larger are characterized are by the amount of elastic tissue contained within their walls, which allows them to both actively vasoconstrict and to change caliber elastically in response to fluctuations in pressure [9, 10]. The pulmonary vascular system is highly dynamic, with many “moving parts” capable of active and passive responses to changes elsewhere in the system. These measurements of blood volume distribution may offer a powerful means of understanding hemodynamic processes more completely and lead to more effective treatments for PVD.

The study has several limitations. All data was collected in the course of clinical practice during a pandemic, and was retrospective, with the result that acquisition protocols varied between centers. The characteristics of the patient were not always available, such that age, gender, and a height could not be well controlled for. Normalization of BVX by total pulmonary blood volume mitigates this somewhat, as previous samplings have shown these measures to vary little across healthy populations.

As noted, areas of opacification confound these algorithms. In most past studies of imaging of pulmonary blood volumes, these opacities have been grounds for excluding scans from analysis. This was neither possible nor desirable in this context given the ubiquity of these findings and their hypothesized role in the pathogenesis of COVID-19 [11-13]. BVX is normalized for total segmented blood volume, which limits the magnitude of error introduced. Notable is the presence of lobes with minimal opacity but markedly reduced BV5 compared to healthy subjects. In combination with the small effect sizes observed in the mixed effect model (Figure 5), this suggests that the detected opacities likely introduce modest error, specifically loss of BV5 and gain of BV10, but are not the singular or even primary driver of these anomalies. We speculate that observed anomalies in vascular volumes are a result of the pathogenic process of the SARS-CoV2 virus itself and possibly the resulting immune response. To the extent that total pulmonary blood volumes are slightly larger in COVID-19 patients than in healthy patients (as was observed in cohorts from Belgium and China), this is accounted for in part, we believe, by the mistaken segmentation of opacities as blood.

Future research will be needed to either better segment areas of opacification or else more completely account for their effect on measurements of pulmonary vascular density. This includes the possibility that, apart from whatever confounding effect they may have, the observed opacities are themselves caused by the same processes that drive BVX anomalies as some researchers have suggested [14]. This understanding is of particular importance in facilitating the use of BVX and BV Spectra in research on PVD secondary to chronic lung disease.

Additional research will seek to improve the accuracy of these methodologies in acquiring the desired measurements in the target populations, as well as to make comparisons with a wealth of clinical data to better understand the part that these hemodynamic redistributions play in the presentation and progression of PVD generally and COVID-19 specifically.

### Informed consent and patient details

The authors declare that this report does not contain any personal information that could lead to the identification of the patient(s). The authors declare that they obtained a written informed consent from the patients and/or volunteers included in the article. The authors also confirm that the personal details of the patients and/or volunteers have been removed.

## Data Availability

Data available upon request

## Disclosure of interest

BL, ML, SB, RG, IK, and JDB are employees of FLUIDDA, a company that develops and markets part of the technology described in this paper. The other authors have no financial relationships with any organization or company that might have an interest in the submitted work and received no direct funding from FLUIDDA.

## Notes

### Funding Statement

No external funding was received.

### Author Declarations

Institutional Review Board approval was granted by the respective local committees from each site which provided patient data: AZ Sint-Maarten (Mechelen), Ziekenhuis Oost-Limburg (Genk), Royal Papworth Hospital, and Wenzhou Medical University. Data for healthy patients comparators were acquired from the COPDGene cohort (NCT00608764).

